# Neural Network Correlates of Apathy, Disinhibition, and Executive Dysfunction in Active-Duty United States Special Operations Forces

**DOI:** 10.1101/2024.05.13.24306944

**Authors:** Natalie Gilmore, Isabella R. McKinney, Chieh-En J. Tseng, Douglas N. Greve, Chiara Maffei, Brian C. Healy, Nicole R. Zürcher, Jacob M. Hooker, Samantha L. Tromly, Daniel P. Perl, Kristen Dams-O’Connor, Christine L. Mac Donald, Brian L. Edlow, Yelena G. Bodien

**Author notes:** **Corresponding Author:** Yelena G. Bodien, 101 Merrimac Street, Suite 310, Boston, MA 02114.

## Abstract

United States Special Operations Forces (SOF) experience neurobehavioral symptoms that can adversely affect training and combat operations. Understanding the neurobiological basis for these symptoms may guide prevention and treatment. In 29 male active-duty SOF with mean (SD) 17(4) years of service, we tested whether self-reported symptoms of apathy, disinhibition, and executive dysfunction measured by the Frontal Systems Behavior Scale, were related to functional magnetic resonance imaging and positron emission tomography biomarkers (translocator protein and tau) of the limbic, salience, and executive control networks. Higher disinhibition was associated with lower functional connectivity and higher tau signal within the salience network.

## Introduction

Special Operations Forces (SOF) are repeatedly exposed to blast overpressure and blunt head trauma during training and combat.^1^ These exposures can lead to neurobehavioral symptoms^2^ that can impact combat performance^3^ and life satisfaction.^4^ There is evidence that neurobehavioral symptoms, including apathy, disinhibition, and executive dysfunction,^5^ are associated with disruption of the large-scale distributed brain networks^6^ supporting these functions. Yet, the neurobiological correlates of the interaction between network dysfunction and neurobehavioral symptoms have not been examined in active-duty SOF, limiting opportunities for brain health optimization and development of targeted interventions.^7^ We examined, in active-duty United States (US) SOF personnel, the relationship between apathy, disinhibition, and executive dysfunction symptoms and functional magnetic resonance imaging (fMRI) as well as positron emission tomography (PET) biomarkers (translocator protein and tau) of the limbic, salience, and executive control networks.

## Methods

We analyzed data collected prospectively as part of the Long-Term Effects of Repeated Blast Exposure in United States Special Operations Forces Personnel (ReBlast) study (ClinicalTrials.gov, NCT05183087). Protocol details have been previously reported.^8^ The ReBlast study was approved by the Mass General Brigham Institutional Review Board (2020P002695) and U.S. Special Operations Command Human Research Protections Office (USSOCOM HRPO; S20010). All participants provided written informed consent.

Participants were male active-duty SOF, age 25 to 45 years, with a history of substantial combat and blast exposure. Over the course of a two-day study visit to Boston, MA, participants completed a comprehensive, multimodal assessment protocol.^8^ Data for the current study included: 1) demographics (e.g., age), military history (e.g., years in service), and exposure to combat (Combat Exposure Scale [CES]^9^); 2) self-reported history of exposure to blast (Generalized Blast Exposure Value [GBEV]^10^) and blunt head trauma (Brain Injury Screening Questionnaire [BISQ]^11^); 3) assessment of neurobehavioral symptom severity (46-item Frontal Systems Behavior Scale [FrSBe]^12^ completed by the SOF participant [FrSBe_Self_] and separately by a designated family member or friend [FrSBe_Family_]); and 4) neuroimaging (structural MRI resting-state fMRI [rs-fMRI], [^11^C]PBR28 translocator protein [TSPO] positron emission tomography [PET], and [^18^F]MK6240 tau PET). Neuroimaging data were acquired on Siemens scanners (Siemens Healthineers, Erlangen, Germany): 1) rs-fMRI data on a 7T Terra MRI scanner with a 32-channel head coil (Nova Medical, Wilmington, MA); and 2) [^11^C]PBR28 TSPO and [^18^F]MK6240 tau PET data on a hybrid PET-MRI scanner with an 8-channel MR head receiver coil (i.e., the BrainPET). Additional information regarding the behavioral and neuroimaging procedures and participant exclusion for specific modalities are included in Supplemental Materials; and procedures have been fully described in our previous work.^8,13^

We conducted univariate (i.e., unadjusted) regression analyses to test for associations between the primary dependent (i.e., FrSBe_Self_ and FrSBe_Family_ Apathy, Disinhibition, and Executive Dysfunction subscale T-scores) and independent (i.e., mean functional connectivity, mean [^11^C]PBR28 TSPO standardized uptake value ratio [SUVR], and mean [^18^F]MK6240 tau SUVR in the limbic, salience, and executive control networks) variables. We also conducted multivariable regression analyses adjusting for age.

## Results

Twenty-nine male participants were mean (SD) 37 (4) years old with mean (SD) 17 (4) years of service. Most participants were enlisted and actively serving in the Army. Combat exposure was moderate-heavy or heavy in 27 out of 29 participants based on the CES. Cumulative blast exposure, as measured by the GBEV, ranged from 387,861 to 363,812,869 with a median value of 9,763,085, which was above a previously-published threshold for symptom reporting.^10^ FrSBe subscale T-scores were above the clinical threshold for impairment (i.e., ≥ 65) for 28% to 31% of participants on the FrSBe_Self_ and 36% to 43% of participants on the FrSBe_Family_. All participants reported exposure to blunt head trauma on the BISQ consistent with a mild TBI, based on Veteran Affairs/Department of Defense criteria.^11^

Results of statistical analyses for FrSBe_Self_ and FrSBe_Family_ Disinhibition Subscale T-scores are presented in Table 2. Higher Disinhibition Subscale T-scores were associated with lower functional connectivity in the salience network on FrSBe_Self_ (β = -0.38, 95% CI [-0.75, -0.005], p = 0.047; Figure 1b) and FrSBe_Family_ (β = -0.37, 95% CI [-0.76, 0.009], p = 0.055). These associations were attenuated after adjusting for age. Higher Disinhibition Subscale T-scores were associated with higher tau signal within the salience network, as measured by [^18^F]MK6240 SUVR, on FrSBe_Self_ (β = 0.38, [0.009, 0.76], p = 0.045, Figure 1), but not FrSBe_Family_ (β = 0.29, [-0.10, 0.69], p = 0.142). FrSBe_Self_ associations with tau signal in the salience network were attenuated after adjusting for age. No associations were observed between FrSBe_Self_ or FrSBe_Family_ Disinhibition Subscale T-scores and TSPO as measured by [^11^C]PBR28 SUVR in the salience network.

**Table 1.**
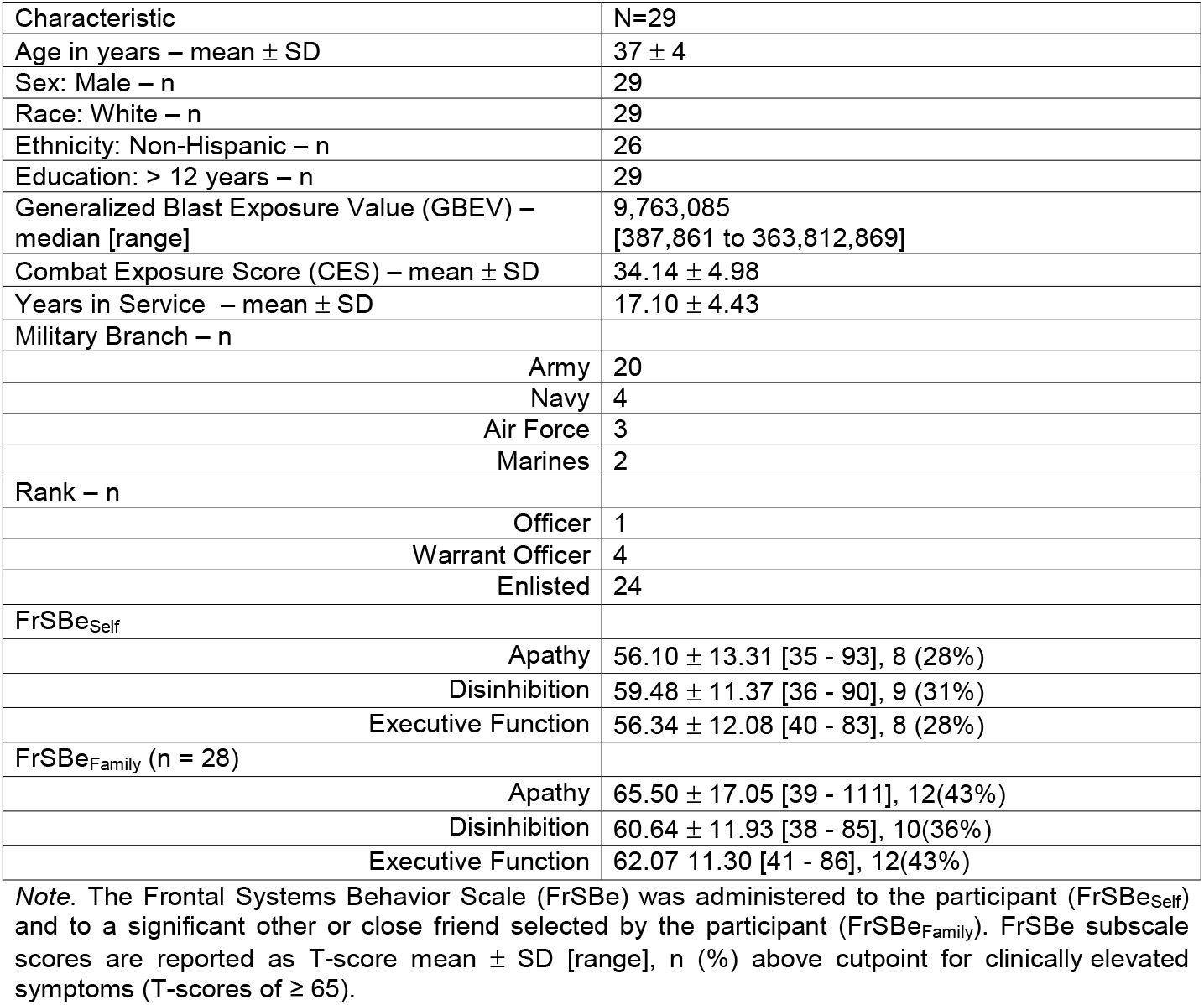
Participant Characteristics.

|  |  |
| --- | --- |
| Characteristic | N=29 |
| Age in years – mean ± SD | 37 ± 4 |
| Sex: Male – n | 29 |
| Race: White – n | 29 |
| Ethnicity: Non-Hispanic – n | 26 |
| Education: > 12 years – n | 29 |
| Generalized Blast Exposure Value (GBEV) – median [range] | 9,763,085<br>[387,861 to 363,812,869] |
| Combat Exposure Score (CES) – mean ± SD | 34.14 ± 4.98 |
| Years in Service – mean ± SD | 17.10 ± 4.43 |
| Military Branch – n |  |
| Army | 20 |
| Navy | 4 |
| Air Force | 3 |
| Marines | 2 |
| Rank – n |  |
| Officer | 1 |
| Warrant Officer | 4 |
| Enlisted | 24 |
| FrSBe<sub>Self</sub> |  |
| Apathy | 56.10 ± 13.31 [35 - 93], 8 (28%) |
| Disinhibition | 59.48 ± 11.37 [36 - 90], 9 (31%) |
| Executive Function | 56.34 ± 12.08 [40 - 83], 8 (28%) |
| FrSBe<sub>Family</sub> (n = 28) |  |
| Apathy | 65.50 ± 17.05 [39 - 111], 12(43%) |
| Disinhibition | 60.64 ± 11.93 [38 - 85], 10(36%) |
| Executive Function | 62.07 ± 11.30 [41 - 86], 12(43%) |

*Note.* The Frontal Systems Behavior Scale (FrSBe) was administered to the participant (FrSBe<sub>Self</sub>) and to a significant other or close friend selected by the participant (FrSBe<sub>Family</sub>). FrSBe subscale scores are reported as T-score mean ± SD [range], n (%) above cutpoint for clinically elevated symptoms (T-scores of ≥ 65).

**Table 2.** Associations between FrSBe_Self_ and FrSBe_Family_ Disinhibition T-scores and the Salience Network.

| Resting State Functional Connectivity |  |  |  |  |  |  |
| --- | --- | --- | --- | --- | --- | --- |
|  | Unadjusted |  |  | Adjusted for Age |  |  |
| | Salience $\beta$ | 95% CI | p | Salience $\beta$ | 95% CI | p |
| Self<br>(n = 28) | -0.378 | -0.751,<br>-0.005 | 0.047 | -0.287 | -0.665,<br>0.091 | 0.13 |
| Family<br>(n = 27) | -0.373 | -0.756,<br>0.009 | 0.055 | -0.304 | -0.705,<br>0.096 | 0.13 |
| [<sup>11</sup>C]PBR28 PET (TSPO signal) SUVR |  |  |  |  |  |  |
|  | Unadjusted |  |  | Adjusted for Age |  |  |
| | Salience $\beta$ | 95% CI | p | Salience $\beta$ | 95% CI | p |
| Self<br>(n = 27) | -0.083 | -0.493,<br>0.328 | 0.681 | -0.177 | -0.561,<br>0.207 | 0.351 |
| Family<br>(n = 26) | 0.046 | -0.374,<br>0.467 | 0.824 | -0.070 | -0.500,<br>0.361 | 0.741 |
| [<sup>18</sup>F]MK6240 PET (tau signal) SUVR |  |  |  |  |  |  |
|  | Unadjusted |  |  | Adjusted for Age |  |  |
| | Salience $\beta$ | 95% CI | p | Salience $\beta$ | 95% CI | p |
| Self<br>(n = 28) | 0.381 | 0.009,<br>0.754 | 0.045 | 0.31 | -0.057,<br>0.676 | 0.094 |
| Family<br>(n = 27) | 0.291 | -0.104,<br>0.685 | 0.142 | 0.223 | -0.165,<br>0.61 | 0.247 |

*Note.* Factors resulting in participants being excluded from Resting State Functional Connectivity and PET analyses are described in Supplementary Methods.
FrSBe = Frontal Systems Behavior Scale; $\beta$ = standardized regression coefficient; TSPO = translocator protein; PET = positron emission tomography, SUVR = standardized uptake value ratio; CI = confidence interval

**Figure 1.**
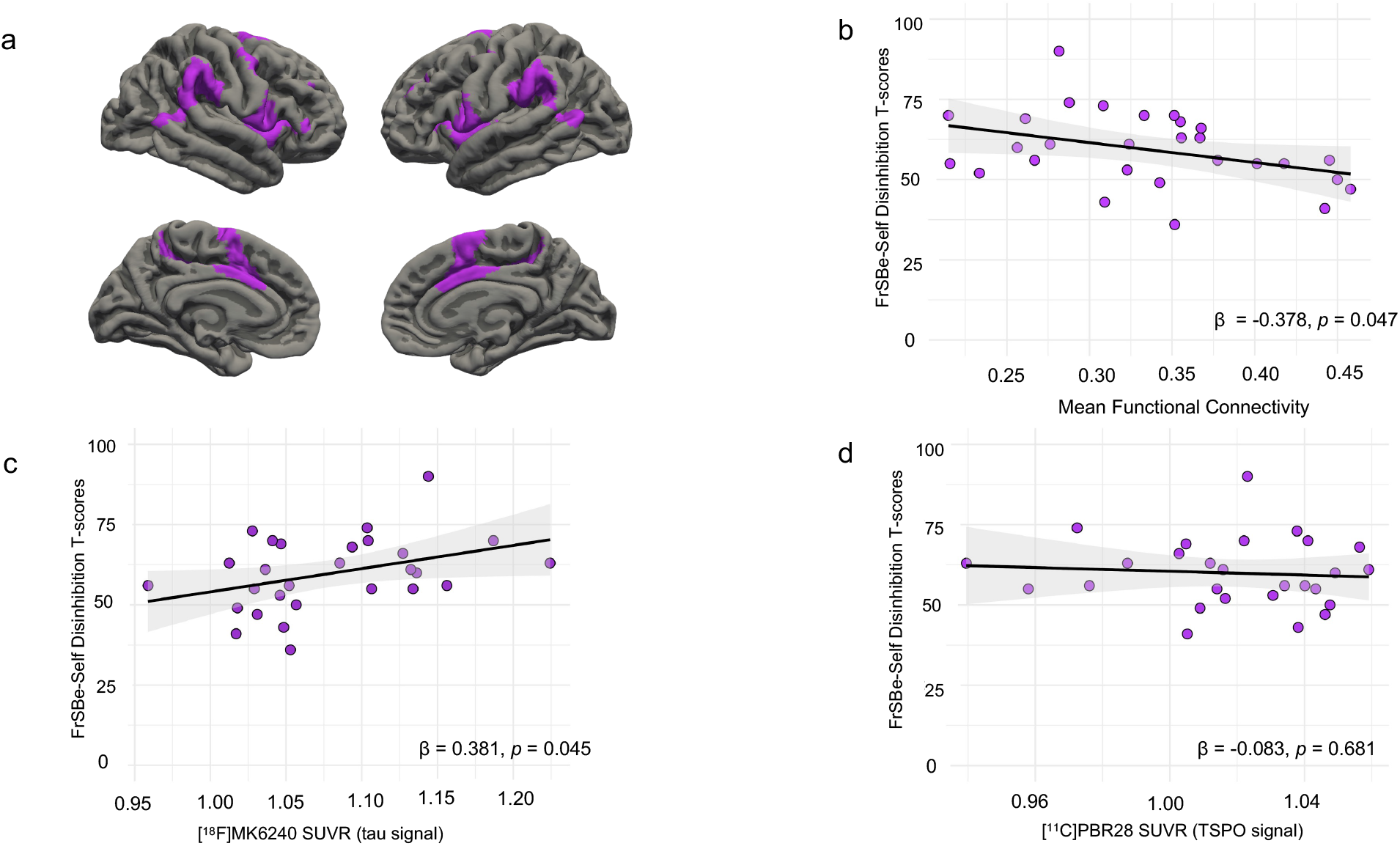
Association of FrSBe_Self_ Disinhibition Scores with Salience Network Biomarkers. The salience network (a, purple regions) was composed of the left inferior frontal gyrus (i.e., pars opercularis), aspects of the left temporal and occipital lobes, aspects of the right temporal, occipital, and parietal lobes, a right precentral region, a right ventral prefrontal region, and bilateral aspects of the frontal lobe including the operculum and insula, the lateral prefrontal, and medial cortices. Higher FrSBe_Self_ Disinhibition T-Scores were associated with lower resting-state functional connectivity (b) and higher tau signal as measured by [^18^F]MK6240 SUVR (c). There was no association between FrSBe_Self_ Disinhibition T-Scores and TSPO signal as measured by [^11^C]PBR28 SUVR (d). Individual data points in b, c, and d are shown as purple circles, regression line is black with the 95% confidence intervals indicated in grey shading. Note: T-scores have a mean of 50 and SD of 10. T-scores ≥ 65 on the FrSBe subscales suggest clinically elevated symptoms. Abbreviations: FrSBe = Frontal Systems Behavior Scale; SUVR = standardized uptake value ratio; TSPO = translocator protein.

Neither FrSBe_Self_ nor FrSBe_Family_ Apathy Subscale T-scores were associated with rs-fMRI or PET biomarkers of the limbic network (Supplemental Table 1). Similarly, neither FrSBe_Self_ nor FrSBe_Family_ Executive Dysfunction Subscale T-scores were associated with rs-fMRI or PET biomarkers of the executive control network (Supplemental Table 2).

## Discussion

In active-duty U.S. SOF with repeated blast exposure, higher self-reported disinhibition symptoms were associated with the salience network across two imaging modalities; we observed similar results with family-reported disinhibition symptoms. The salience network is a known contributor to cognitive and emotional dysregulation, and its structural as well as functional disruption has been linked to neuropsychiatric disorders for which disinhibition is a hallmark characteristic.^14^

Our findings build on recent work in the same cohort of active-duty SOF wherein higher blast exposure was related to higher cortical thickness in bilateral rostral anterior cingulate cortex (rACC) and right insula, and lower TSPO signal in the right rACC, all of which are regions within the salience network.^13^ Interestingly, in a different cohort of active-duty SOF, higher blast exposure was associated with increased TSPO signal in the salience network.^15^ The discrepancy in findings across these two studies^13,15^ may have been due to the use of a different tracer (i.e., [18F]DPA-714), a different statistical analytic approach (i.e., applying dimensionality reduction before testing the association between biomarkers and blast exposure), or a different study design (i.e., comparing blast-exposed special operators to non-exposed matched controls). Overall, the present work illustrates the potential neurobehavioral consequences of network-level disruptions in a population heavily exposed to blast and blunt head trauma.

Approximately one-third of participants and their family members reported apathy, disinhibition and executive function symptoms that exceed the clinical threshold for impairment (Table 1). This result is consistent with prior studies showing that a substantial proportion of service members with mild TBI,^16^ including SOF,^2^ experience neurobehavioral symptoms. Targeted measures aimed at preventing or treating salience network disruption may promote quality of life for SOF and their families.^4^

Adjusting our statistical models for age attenuated the association between disinhibition and functional connectivity as well as tau signal in the salience network, which supports prior literature showing that disinhibition increases with age.^17^ Symptoms of apathy and executive control were not related to neuroimaging biomarkers of the limbic or executive control networks, respectively. The Yeo 7-network atlas,^18^ which we used to identify regions of interest, only includes cortical regions. Consequently, the contribution of key subcortical regions could not be assessed (e.g., hippocampus and amygdala within the limbic network; thalamus and ventral striatum within the salience network).^14^ Furthermore, the neuroanatomical substrates of executive control are highly distributed and poorly defined,^19^ potentially contributing to the lack of an association between executive dysfunction symptoms and biomarkers of the executive control network. There was no association between TSPO signal and disinhibition. However, higher tau signal was associated with higher symptoms of disinhibition. While the MK-6240 ligand binds to tau pathology,^20^ and thus the association between higher tau signal in the salience network and higher disinhibition symptoms may represent neurofibrillary tangles, it could also represent dystrophic neurites and other pathological cellular processes that can only be confirmed by post-mortem examination of the brain. The study was limited by lack of longitudinal data and a small sample size. Nevertheless, the findings contribute to our understanding of the neural network correlates of disinhibition in active-duty SOF.

## Conclusions

Higher self-reported symptoms of disinhibition in active-duty SOF with heavy exposure to blast and blunt head trauma were associated with lower functional connectivity and higher tau signal in the salience network, which is known to contribute to emotional and cognitive function. These findings support the use of a multimodal, network-based approach to examine the effects of head trauma on active-duty SOF brain health and highlight a potential role for the salience network in the prevention and treatment of neurobehavioral symptoms in this population.

## Data Availability

USSOCOM policy limits public sharing of data generated and analyzed for the current paper. Any future requests for these data may be submitted to the corresponding author for subsequent vetting and approval by USSOCOM.

## Abbreviations

SOF: Special Operations Forces
ReBlast: Long-Term Effects of Repeated Blast Exposure in United States Special Operations Forces Personnel
GBEV: Generalized Blast Exposure Value
BISQ: Brain Injury Screening Questionnaire
FrSBe: Frontal Systems Behavior Scale
FrSBe_Self_: FrSBe completed by SOF participant
FrSBe_Family_: FrSBe completed by designated family member or friend of SOF participant
rs-fMRI: resting-state functional MRI
TSPO: translocator protein
PET: positron emission tomography
SUVR: standardized uptake value ratio
rACC: rostral anterior cingulate cortex
TBI: traumatic brain injury
USSOCOM: United States Special Operations Command

## Acknowledgments

We thank the participants and their families for their time, candor, and meaningful contributions to this study. We appreciate Katryna Deary and Jessica Kelemen for their assistance in recruitment and screening participants. We value the contributions of the ReBlast MRI Safety Committee (John E. Kirsch, Jacob C. Calkins, Amy L. Kendall, and Grae E. Arabasz) and the regulatory and grants administrative support staff (Jennifer Michaud, Kelsey Radmanesh, Maryam Masood). We also appreciate all those who were involved in MRI (Gabriel Llorden Ramos, Kyle Droppa) and PET data acquisition (Mary Tresvalles, Mariah Manter, Kyle Droppa, Camila Canales, Riana Schleicher, Shirley Hsu, Oliver Ramsay, Alison Brown, Anne Siewko), including PETNET Solutions New York for radiotracer production. This work was funded by the U.S. Department of Defense (USSOCOM Contract No. H9240520D0001) and the Navy SEAL Foundation.

## Author Contributions

Author contributions included conception and study design (N.G., I. R. M., N.R.Z, J. M. H., B.C.H., B.L.E, Y.G.B.), data collection or acquisition (N.G., C.J.T., C.M., D.N.G), statistical analysis (N.G., B.C.H.), interpretation of results (N.G., B.C.H., B.L.E., Y.G.B.), drafting the manuscript work or revising it critically for important intellectual content (N.G., I.R.M, C.J.T., D.N.G., C.M., S.L.T., D.P.P., K.D.P., C.L.M., B.L.E., Y.G.B.) and approval of final version to be published and agreement to be accountable for the integrity and accuracy of all aspects of the work (All authors).

## Disclaimer

The opinions and assertions expressed herein are those of the author(s) and do not reflect the official policy or position of the Uniformed Services University of the Health Sciences or the Department of Defense.

## Potential Conflicts of Interest

The authors report no conflicts of interest.

## Notes

### Competing Interest Statement

The authors have declared no competing interest.

### Clinical Protocols

https://doi.org/10.1089/neu.2022.0030

### Author Declarations

The ReBlast study was approved by the Mass General Brigham Institutional Review Board (2020P002695) and U.S. Special Operations Command Human Research Protections Office (USSOCOM HRPO; S20010).

