## Supplemental Materials for "Neural Network Correlates of Apathy, Disinhibition, and Executive Dysfunction in Active-Duty United States Special Operations Forces"

Natalie Gilmore[1,2], Isabella R. McKinney[1,2], Chieh-En J. Tseng[3], Douglas N. Greve[3], Chiara Maffei[1,2,3], Brian C. Healy[2,4], Nicole R. Zürcher[3], Jacob M. Hooker[3], Samantha L. Tromly[5], Daniel P. Perl[6], Kristen Dams-O'Connor[7], Christine L. Mac Donald[8], Brian L. Edlow MD[1,2,3], Yelena G. Bodien[1,2,9]

**Author Affiliations:**
[1]Center for Neurotechnology and Neurorecovery, Massachusetts General Hospital, Boston, MA 02114
[2]Department of Neurology, Massachusetts General Hospital and Harvard Medical School, Boston, MA 02114
[3]Department of Radiology, Athinoula A. Martinos Center for Biomedical Imaging, Massachusetts General Hospital and Harvard Medical School, Charlestown, MA 02129
[4]Biostatistics Center, Massachusetts General Hospital, Boston, MA 02114
[5]Institute of Applied Engineering, University of South Florida, Tampa, FL 33612
[6]Department of Pathology, F. Edward Hébert School of Medicine, Uniformed Services University (USU), Bethesda, Maryland 20814, United States
[7]Department of Rehabilitation and Human Performance, Icahn School of Medicine at Mount Sinai, New York, NY 10029
[8]Department of Neurological Surgery, University of Washington, Seattle, WA 98195
[9]Department of Physical Medicine and Rehabilitation, Spaulding Rehabilitation Hospital and Harvard Medical School, Charlestown, MA 02129

**Table of Contents**


| | |
|---|---|
| Supplemental Text: Methods | 2 |
| Supplemental Table 1. Associations between $\text{FrSBe}_{\text{Self}}$ and $\text{FrSBe}_{\text{Family}}$ Apathy T-scores and the Limbic Network | 6 |
| Supplemental Table 2. Associations between $\text{FrSBe}_{\text{Self}}$ and $\text{FrSBe}_{\text{Family}}$ Executive Dysfunction T-scores and the Executive Control Network | 7 |


**Abbreviations:**
BISQ = Brain Injury Screening Questionnaire
PET-MRI = positron emission tomography-magnetic resonance imaging
ROI = regions of interest
rs-fMRI = resting state functional magnetic resonance imaging
BOLD = blood-oxygen-level-dependent

**Supplemental Text: Methods**

*Behavioral Data Processing*

Exposures: We administered the Brain Injury Screening Questionnaire (BISQ) to capture lifetime history of traumatic brain injury (TBI). As part of the BISQ, we queried whether participants had experienced a blow to the head in at least 20 different situations, including combative training and contact sports, and if so, how many times. Although the BISQ asks about the duration of dazed and confused symptoms and loss of consciousness for all occurrences, we only asked for additional detail regarding the three worst occurrences as many participants could not recall a discrete number of events. As described in the main text, all participants reported at least one blow to the head consistent with mild TBI according to VA/DoD guidelines. Finally, we administered the Generalized Blast Exposure Value (GBEV) to capture lifetime blast exposure. We log-transformed the GBEV to account for the wide range of scores (see Table 1) and facilitate visualization.

Neurobehavioral Symptoms: We summed the item ratings to obtain Apathy, Disinhibition, and Executive Function subscale scores and a Total score. We did not use the total score as we were interested in testing hypothesized relationships between specific symptoms and networks (e.g., Apathy and the Limbic Network; Disinhibition and the Salience Network; Executive Dysfunction and the Executive Control Network). To improve interpretability and allow for comparison across subscales, we followed procedures described in the manual to convert raw FrSBe subscale scores to T-scores using age-, sex-, and education-level reference data.

Participants were asked to suggest a family member or close other that could complete the $FrSBe_{Family}$. $FrSBe_{Family}$ respondents were significant others, either current or former, except for one participant who was unpartnered at the time and suggested a close friend. One participant declined to provide a family member/close other for completion of the $FrSBe_{Family}$ and was excluded from all analyses using the $FrSBe_{Family}$ subscales.

The FrSBe inquires about symptoms “before the illness or injury” and “at the present time.” We asked ReBlast participants to report on their symptoms “before enlistment” because there was no index illness or injury that prompted study inclusion. There was variability in the timespan between enlistment and study participation and not all $FrSBe_{Family}$ respondents knew participants before enlistment. Thus, we assessed the FrSBe data “at the present time” only.

*Neuroimaging Data Processing*

Structural magnetic resonance imaging (MRI): We processed the T1-weighted images using Freesufer’s “recon-all” pipeline, providing cortical parcellations and subcortical segmentations for each individual subject that we used to analyze the positron emission tomography-MRI (PET-MRI) and resting state functional MRI (rs-fMRI) data.

PET-MRI: The PET emission data for translocator protein (TSPO; 60 to 90-minute post-radioligand injection) and tau (70 to 90-minute post-radioligand injection) were divided into five-minute frames. We reconstructed the standardized uptake value (SUV) images using an MRI-based attenuation map, then, re-aligned and averaged them. We linearly registered the SUV to each participant's structural scan using FreeSurfer. We normalized SUV to a pseudo-reference region (i.e., calculated SUVR; TSPO: whole brain, excluding the ventricles; tau: isthmus cingulate cortex). We transformed each participant's T1-weighted MRI from native space to MNI space using FSL. We applied the inverse-transformation to the Yeo-7 network atlas, mapped the atlas to each participant's native space, and extracted native space SUVRs. Three participants with low affinity binding for [$^{11}$C]PBR28 were excluded from the TSPO PET-MRI analyses. Two participants who did not receive tau PET scans due to [$^{18}$F]MK6240 production failures were excluded from the tau PET-MRI analyses.

Resting state functional magnetic resonance imaging (rs-fMRI): We processed these data in FreeSurfer 7.3.0. We mapped the Yeo-7 network atlas to each participant's surface. We projected the atlas out of the surface into the T1 anatomical volume for each participant. We mapped the T1 anatomical volume into the rs-fMRI space. We used the split components version of the Yeo-7 network atlas (https://github.com/ThomasYeoLab/CBIG/blob/master/stable_projects/brain_parcellation/Yeo2011_fcMRI_clustering/1000subjects_reference/Yeo_JNeurophysiol11_SplitLabels/project_to_individual/Yeo2011_7networks_Split_Components_LUT.txt). Regions of interest (ROI) with bilateral representation in the atlas were merged to create a single ROI for that area. There were 8 ROIs for the salience network: left pars opercularis (ParOper), left temporal occipital (TempOcc), right temporal-occipital-parietal (TempOccPar), right precentral (PrC), right ventral prefrontal (PFCv), and bilateral frontal-operculum-insula (FrOperIns), lateral prefrontal (PFCl), and medial (Med) cortex. There were 9 ROIs for the executive control network: left dorsal prefrontal (PFCd), left OFC, and bilateral parietal (Par), temporal (Temp), lateral prefrontal cortex (PFCl), ventral prefrontal cortex (PFCv), precuneus (pCun), cingulate (Cing), and medial prefrontal (Med) cortices. There were 2 regions of interest for the limbic network: bilateral orbitofrontal cortex (OFC) and temporal pole (TempPole). We extracted the average blood-oxygen-level-dependent (BOLD) signal for each ROI. We tested the association between the average BOLD signal in each ROI with every other ROI in the network. We computed the Pearson correlation coefficient between each ROI pair within each network after regressing out the effects of motion, white matter signal, and CSF signal. We averaged the Pearson correlation coefficients for each ROI pair in the network to obtain the mean functional connectivity for the network. Two participants were excluded from the rs-fMRI analyses due to signal inhomogeneities.

Supplemental Table 1. Associations between $FrSBe_{Self}$ and $FrSBe_{Family}$ Apathy T-scores and the Limbic Network

| Resting State Functional Connectivity | | | | | | |
|---|---|---|---|---|---|---|
| | Unadjusted | | | Adjusted for Age | | |
| | Limbic β | 95% CI | p | Limbic β | 95% CI | p |
| Self (n = 28) | 0.004 | -0.399, 0.407 | 0.984 | -0.009 | -0.434, 0.417 | 0.967 |
| Family (n = 27) | 0.088 | -0.322, 0.499 | 0.662 | -0.008 | -0.403, 0.386 | 0.966 |
| [11C]PBR28 (TSPO signal) PET SUVR | | | | | | |
| | Unadjusted | | | Adjusted for Age | | |
| | Limbic β | 95% CI | p | Limbic β | 95% CI | p |
| Self (n = 27) | -0.011 | -0.423, 0.401 | 0.957 | -0.02 | -0.449, 0.409 | 0.924 |
| Family (n = 26) | 0.242 | -0.167, 0.651 | 0.234 | 0.171 | -0.215, 0.557 | 0.368 |
| [18F]MK6240 PET (tau signal) SUVR | | | | | | |
| | Unadjusted | | | Adjusted for Age | | |
| | Limbic β | 95% CI | p | Limbic β | 95% CI | p |
| Self (n = 28) | -0.223 | -0.616, 0.17 | 0.253 | -0.222 | -0.625, 0.181 | 0.267 |
| Family (n = 27) | -0.020 | -0.432, 0.392 | 0.921 | 0.009 | -0.376, 0.395 | 0.960 |

*Note.* FrSBe = Frontal Systems Behavior Scale; β = standardized regression coefficient; TSPO = translocator protein; PET = positron emission tomography, SUVR = standardized uptake value ratio; CI = confidence interval

Supplemental Table 2. Associations between $FrSBe_{Self}$ and $FrSBe_{Family}$ Executive Dysfunction T-scores and the Executive Control Network

| Resting State Functional Connectivity | | | | | | |
|---|---|---|---|---|---|---|
| | Unadjusted | | | Adjusted for Age | | |
| | Executive Control β | 95% CI | p | Executive Control β | 95% CI | p |
| Self (n = 28) | -0.001 | -0.404, 0.402 | 0.997 | 0.053 | -0.389, 0.494 | 0.808 |
| Family (n = 27) | 0.209 | -0.194, 0.612 | 0.296 | 0.216 | -0.230, 0.663 | 0.327 |
| [11C]PBR28 (TSPO signal) PET SUVR | | | | | | |
| | Unadjusted | | | Adjusted for Age | | |
| | Executive Control β | 95% CI | p | Executive Control β | 95% CI | p |
| Self (n = 27) | -0.173 | -0.578, 0.233 | 0.389 | -0.238 | -0.666, 0.190 | 0.263 |
| Family (n = 26) | 0.079 | -0.341, 0.499 | 0.700 | 0.117 | -0.335, 0.57 | 0.597 |
| [18F]MK6240 (tau signal) PET SUVR | | | | | | |
| | Unadjusted | | | Adjusted for Age | | |
| | Executive Control β | 95% CI | p | Executive Control β | 95% CI | p |
| Self (n = 28) | -0.123 | -0.523, 0.277 | 0.533 | -0.141 | -0.548, 0.266 | 0.483 |
| Family (n = 27) | 0.045 | -0.367, 0.456 | 0.824 | 0.049 | -0.374, 0.471 | 0.815 |

*Note.* FrSBe = Frontal Systems Behavior Scale; β = standardized regression coefficient; TSPO = translocator protein; PET = positron emission tomography, SUVR = standardized uptake value ratio; CI = confidence interval
